# The Impact of Normal Aging Out on Continuity of Treatment among Children and Adolescents Living with HIV in the PEPFAR Supported Program in Kenya

**DOI:** 10.1101/2024.01.16.24301354

**Authors:** Lydia Odero, Aaron Chafetz, Mary Gikura, Deborah Goldstein, Rachel Golin, Diana Kemunto, Nelly Maina, Immaculate Mutisya, Kennedy Muthoka, Evelyn Nganga, Tishina Okegbe, Salome Okutoyi, Gonza Omoro, George Siberry, Rose Wafula, Dunstan Achwoka

**Affiliations:** United States Agency for International Development (USAID), Nairobi, Kenya; Office of HIV/AIDS, United States Agency for International Development (USAID), Washington, DC, USA; Palladium Kenya, Nairobi, Kenya; GHTASC, Credence Management Solutions LLC, supporting United States Agency for International Development (USAID), Office of HIV/AIDS, Washington, DC, USA; U.S. Centres for Disease Control (CDC), Nairobi, Kenya; U.S. Department of Defense, Nairobi, Kenya; National AIDS and STI Control Programme (NASCOP), Nairobi, Kenya

**Keywords:** Adolescents, Aging-out, HIV, HIV care continuum, Care and Treatment, Pediatrics

## Abstract

**Introduction:** To reduce HIV-related morbidity and mortality among children and adolescents living with HIV, defined as those 0-14 years old, continuity of treatment is critical. Treatment continuity estimates among children and adolescents living with HIV lag adults. We sought to understand how aging out among children and adolescents living with HIV in Kenya impacts continuity of treatment.

**Methods:** A retrospective cohort analysis was performed on de-identified individual-level data from the Kenya National Data Warehouse for all clients who initiated and/or received antiretroviral therapy between the periods of Oct 1, 2018 and Sep 30, 2022 (US Government fiscal years 2019-2022). Children and adolescents living with HIV previously on antiretroviral therapy and those newly initiating antiretroviral therapy were included in the analysis. Outcomes included aging out of the cohort after turning 15 years old, interruption in treatment, return to treatment, and remaining active on treatment.

**Results:** The study analyzed client-level data for four US Government fiscal years 2019-2022. The number of active children and adolescents living with HIV on treatment at the end of fiscal year 2019 was 44,628. This changed to 48,218, 48,262, and 44,780 representing 8%, 0%, and -7% cohort growth/loss at the end of fiscal years 2020, 2021 and 2022, respectively. Among those who were on treatment at the beginning of each fiscal year, aging-out accounted for 47%, 39%, and 28% of the total losses for the periods 2020, 2021 and 2022, respectively. Interruptions in treatment accounted for proportions ranging from 5-9% among those active on treatment. Among the newly-initiated on treatment within each fiscal year, aging-out proportions ranged from 3%-5%. Among those who returned to treatment in each fiscal year, the proportions who remained active at the end of the fiscal year varied from 72%-76%.

**Conclusions:** This analysis suggests that normal aging-out results in underestimation of HIV treatment continuity for children and adolescents living with HIV. Routine aging out analyses can inform programs on their true rates of interruptions in treatment, as they work to achieve epidemic control among children and adolescents living with HIV.

## Introduction

Despite the “Treat All” era, pediatric antiretroviral therapy (ART) coverage remains well-below adult coverage, with 63% ART coverage rates for children and adolescents living with HIV (C/ALHIV) (0-14 years) in Eastern and Southern Africa compared to 83% for adults (≥15 years) [1]. Only 70% of C/ALHIV in the region have a known HIV status, and of those receiving ART, only 51% are virally suppressed [2]. These dire statistics are attributed in part to inadequate antenatal care to prevent vertical transmission, poor retention in care among pregnant women with HIV, and the rate of HIV infection among children while breastfeeding [3]. Poor viral load suppression (VLS) and interruptions in treatment (IIT) among C/ALHIV has been associated with among others, difficult to take ART formulations; history of opportunistic infections; and caregiver’s VLS status and household vulnerability [4-8].

Optimized case-finding and enhanced linkage to treatment are needed to close gaps in ART coverage and HIV-related morbidity and mortality for C/ALHIV [9]. While HIV continuity of treatment (CoT) is essential for VLS, prevention of disease progression and mortality, and further transmission; data is limited on impact of aging on CoT proxies used for programmatic evaluation and improvement.

Tanzanian program data revealed substantial loss in the number of C/ALHIV on ART in 2015. Subsequently, systematic retrospective analysis led to development of a standardized tool for loss analyses for those aged 0-14 years (unpublished program data). This tool was refined in 2019, when t<u>he U.S. President’s Emergency Plan for AIDS Relief</u> (PEPFAR) instituted IIT reporting. Included were C/ALHIV who had: -transferred out of a clinical site; refused or stopped ART; experienced IIT; and died [10]. These site-level data provided granular assessment of C/ALHIV experiencing IIT and were used to optimize CoT and achieve VLS.

In 2021, PEPFAR advised programs to routinely review adolescent fine age bands to identify ongoing gaps in CoT and VLS [11]. Since 2022, PEPFAR guidance has encouraged programs to evaluate electronic medical records (EMRs) and person-based registries to assess actual experience of aging out in pediatric cohorts [12-13]. However, because PEPFAR’s data is reported at aggregate and not individual-level, analysis of CoT follows the same and does not correctly account for clients who ‘age out’ during transitions between fiscal years (FY).

Analysis of PEPFAR data from 2018-2019 in eight African countries showed that aging can distort age-band retention estimates [14]. This study used 1-year age-bands from 2018 SPECTRUM modeling data to adjust for aging and reported that the adjustment increased ART retention estimates for those aged <1 and 1-9 years but had variable effects on retention for 10-14 and 15-19 years. Authors recommended country-specific analysis to better understand implications of ‘aging out’ for these age bands and better optimize C/ALHIV CoT [14]. In Tanzania, the USAID Afya Yangu Northern project reported that in FY2022, aging out contributed to 48% of the loss in current on ART among C/ALHIV and adjustment for aging out improved pediatric proxy retention from 86% to 96% and program growth from -2% to 10% (unpublished data). Additionally, retrospective cohort analysis of C/ALHIV on ART in Kenyan USAID-supported facilities from 2019-2020 showed that normal aging results in underestimated CoT rates [15].

Compared to aggregate-level data, individual-level data presents a better platform to describe the aging effect among clients on treatment. Evolution of EMRs and establishment of the Kenya National Data Warehouse (NDW) provides such an opportunity [16]. This analysis utilizes individual-level data from the NDW to understand how C/ALHIV aging-out impacts CoT in Kenya.

## Methods

### Study Design

The study employed a retrospective cohort observational design to investigate aging out and treatment interruptions among C/ALHIV on treatment. The analysis included all clients 0-14 years old at any time period between October 1, 2018, and September 30, 2022 [FY2019 – FY2022)] and were either newly initiated on ART or currently on ART during the same period.

### Materials or Instruments & Data Collection

De-identified client records were accessed from the National Data Warehouse (NDW) on April 18, 2023, and extracted for analysis. NDW is a repository of de-identified individual-level client data offered care and treatment services at health facilities. Facility-based health-care workers use the Ministry of Health comprehensive care clinic card (MOH 257) to document clinical notes for clients either newly initiating ART or attending subsequent clinical visits. This tool/card is digitized in the facility EMR system. De-identified data from the facility EMR is then transmitted to the NDW monthly. The authors did not have information that could identify individual clients during or after data collection. RStudio version 2022.12.0 was used for the analysis.

### Variables

Data were extracted from the NDW using the following variables: facility name, master facility listing code, client ID, date of birth, sex, ART initiation date, ART regimen at initiation, current ART regimen, appointment and visit dates. Using client’s appointment and visit dates, the outcomes below were computed per FY:

1. Aged out - a client who turns 15 years old before or at the end of FY.
2. IIT - a client with no clinical contact for 28 days after their last scheduled appointment or expected clinical contact.
3. Return to treatment - a client who experienced IIT during any previous FYs, who successfully reinitiated ART within the FY of interest and remained on treatment until the end of that FY.
4. Active on treatment - a client who is currently receiving ART within a FY and remained on ART as at the end of the FY.
5. Unclassified - a category for clients who do not fit into any of the defined categories mentioned above.

### Data Analysis

Data analysis involved a multi-step approach to longitudinal data on clients’ status across multiple FYs with data cleaning and quality control procedures to eliminate irrelevant or incomplete records. The dataset was then grouped according to client identifiers and FYs; and aggregation techniques were employed to calculate counts of clients into various outcome categories for each FY. Descriptive techniques were used to summarize and present the data’s key features. A Sankey diagram was used to present visualization of clients’ movement from one outcome category at the beginning or in the course of a given FY to another category at the end of the same FY.

### Ethical Considerations

The research was approved by Jomo Kenyatta University of Agriculture and Technology, Ethics and Research Committee (JKU/ISERC/02316/0880); Office of the Associate Director for Science, Centers for Disease Control and Prevention (CDC) with Accession #: CGH-KEN-1/18/23-8faf7; and research license obtained from the National Commission for Science, Technology & Innovation (NACOSTI/P/23/27455). Client’s consent was not required as the authors neither interacted with the clients nor had access to identifiable data.

## Results

Clinical records of C/ALHIV on treatment or initiating treatment in the Kenyan NDW were analyzed for a period of four FYs 2019-2022. At the end of FY2019, 44,628 (72.7%) C/ALHIV remained on treatment. At the end of each of the FYs 2020, 2021, and 2022, 48,218 (78.6%), 48,262 (78.7%) and 44,780 (73.0%) C/ALHIV remained active on treatment (Table 1).

**Table 1:** Kenya Children and Adolescent HIV Treatment Cohort, National Data Warehouse, Fiscal Years (FY) 2020 - 2022.

|  | **FY2020**<br>**(n= 61,357)** | **FY2021**<br>**(n=60,610)** | **FY2022**<br>**(n=55,166)** |
| --- | --- | --- | --- |
| **Status During Fiscal Year** |  |  |  |
| *Newly initiating ART* | 5,183 (8.4%) | 3,264 (5.4%) | 2,047 (3.7%) |
| *Returned to Treatment* | 3,841 (6.3%) | 3,191 (5.3%) | 2,118 (3.8%) |
| *Unclassified* | 7,705 (12.6%) | 5,942 (9.8%) | 2,752 (5.0%) |
| *Current on Treatment (prior fiscal year)* | 44,628 (72.7%) | 48,213 (79.5%) | 48,249 (87.5%) |
| **Status End of Fiscal Year** |  |  |  |
| *Active on Treatment* | 48,218 (78.6%) | 48,262 (78.7%) | 44,780 (73.0%) |
| *Interruptions in Treatment* | 6,903 (11.3%) | 5,430 (8.8%) | 3,400 (5.5%) |
| *Aged Out* | 5,603 (9.1%) | 6,734 (11.0%) | 6,986 (11.4%) |
| *Unclassified* | 633 (1.0%) | 184 (0.3%) | - (0.0%) |
| **Loss/Gain (%)** | 3,590(8%) | 49(0%) | -3,469 (-7%) |

Overall, the cohort grew by 3,590 (8%) in FY2020 and reduced by 3,469 (7%) by the end of FY 2022. C/ALHIV who had disengaged from care and returned to treatment captured within the course of FYs 2020, 2021, and 2022 stood at 3,841 (6.3%), 3,191 (5.3%), and 2,118 (3.8%) respectively. The cohort of C/ALHIV interrupting treatment at the end of FY2020, FY2021 and FY2022 were 6,903 (11.3%), 5,430 (8.8%) and 3,400 (5.5%) while 5,603 (9.1%), 6,734 (11.0%) and 6,986 (11.4%) C/ALHIV aged out at the end of each of the respective FYs (Table 1).

Over the four FYs from 2019-2022, 80% of C/ALHIV remained active on treatment among the current on treatment cohort (i.e. those who were already on treatment at the start of the FY). For this cohort, IIT stood at 4,187 (9%), 3,864 (8%), and 2,564 (5%) at the end of each of the three FY transitions. In FY2020, 4,698 (11%) of the current on treatment cohort aged out, while in FY2021 and FY2022, 5,944 (12%) and 6,489 (13%) of the cohort aged out. IIT accounted for 53%, 61% and 72% of the total losses experienced in FY2020, FY2021 and FY2022 respectively. Losses attributed to aging out among clients currently on treatment were 47% in FY2020, 39% in FY21, and 28% in FY2022 (Table 2).

**Table 2:** Kenya C/ALHIV Aging Out Analysis (at end of FY2020, 2021 and 2022)

|  | **FY2020** | **FY2021** | **FY2022** |
| --- | --- | --- | --- |
| Among Active on Treatment (prior fiscal year) | **(n= 44,628)** | **(n= 48,213)** | **(n= 48,249)** |
| *Remained Active on Treatment (%)* | 35,743 (80%) | 38,405 (80%) | 39,196 (81%) |
| *Interruptions in Treatment (IIT) (%)* | 4,187 (9%) | 3,864 (8%) | 2,564 (5%) |
| *Aged Out (%)* | 4,698 (11%) | 5,944 (12%) | 6,489 (13%) |
| *% Loss attributed to aging out* | 47% | 39% | 28% |
| *% Loss attributed to IIT* | 53% | 61% | 72% |
| Among Clients Newly Initiated on Treatment (current fiscal year) | **(n= 5,183)** | **(n= 3,264)** | **(n=2,047)** |
| *Remained Active on Treatment (%)* | 3,353 (65%) | 2,534 (78%) | 1,721 (84%) |
| *Interruptions in Treatment (IIT) (%)* | 1,048 (20%) | 429 (13%) | 233 (11%) |
| *Aged Out (%)* | 149 (3%) | 117 (4%) | 93 (5%) |
| *Unclassified* | 633 | 184 |  |
| Returned to Treatment | **(n=3,841)** | **(n=3,191)** | **(n=2,118)** |
| *Remained Active on Treatment (%)* | 2,904 (76%) | 2,294 (72%) | 1,575 (74%) |
| *Interruptions in Treatment (IIT) (%)* | 647 (17%) | 646 (20%) | 352 (17%) |
| *Aged Out (%)* | 290 (8%) | 251 (8%) | 191 (9%) |

Among C/ALHIV who newly initiated ART, 3,353 (65%), 2,534 (78%), and 1,721 (84%) remained active across the three FY transitions into FY2020, FY2021, and FY2022, respectively. IIT in this cohort stood at 1,048 (20%), 429 (13%), and 233 (11%) across the three FYs. Aging out among C/ALHIV who newly initiated ART consisted of 149 (3%), 117 (4%) and 93 (5%) clients across the three FY transitions.

Among the cohort of C/ALHIV who returned to treatment, those who remained active till end of the FY consisted of 2,904 (76%), 2,294 (72%) and 1,575 (74%) across FY2020, FY2021 and FY2022 respectively. IIT were 647 (17%), 646 (20%), 352 (17%) while aging out was 290 (8%), 251 (8%) and 191 (9%) for the three FYs (Table 2).

In 2020, 7,705 clients were in the unclassified category, making up 8% of the total. However, by the end of 2020, this number dropped to 633 (92% decrease).

The movement of clients from one category to the other between fiscal years is depicted by the Sankey diagrams as shown in Figs 1-3.

**Fig 1.**
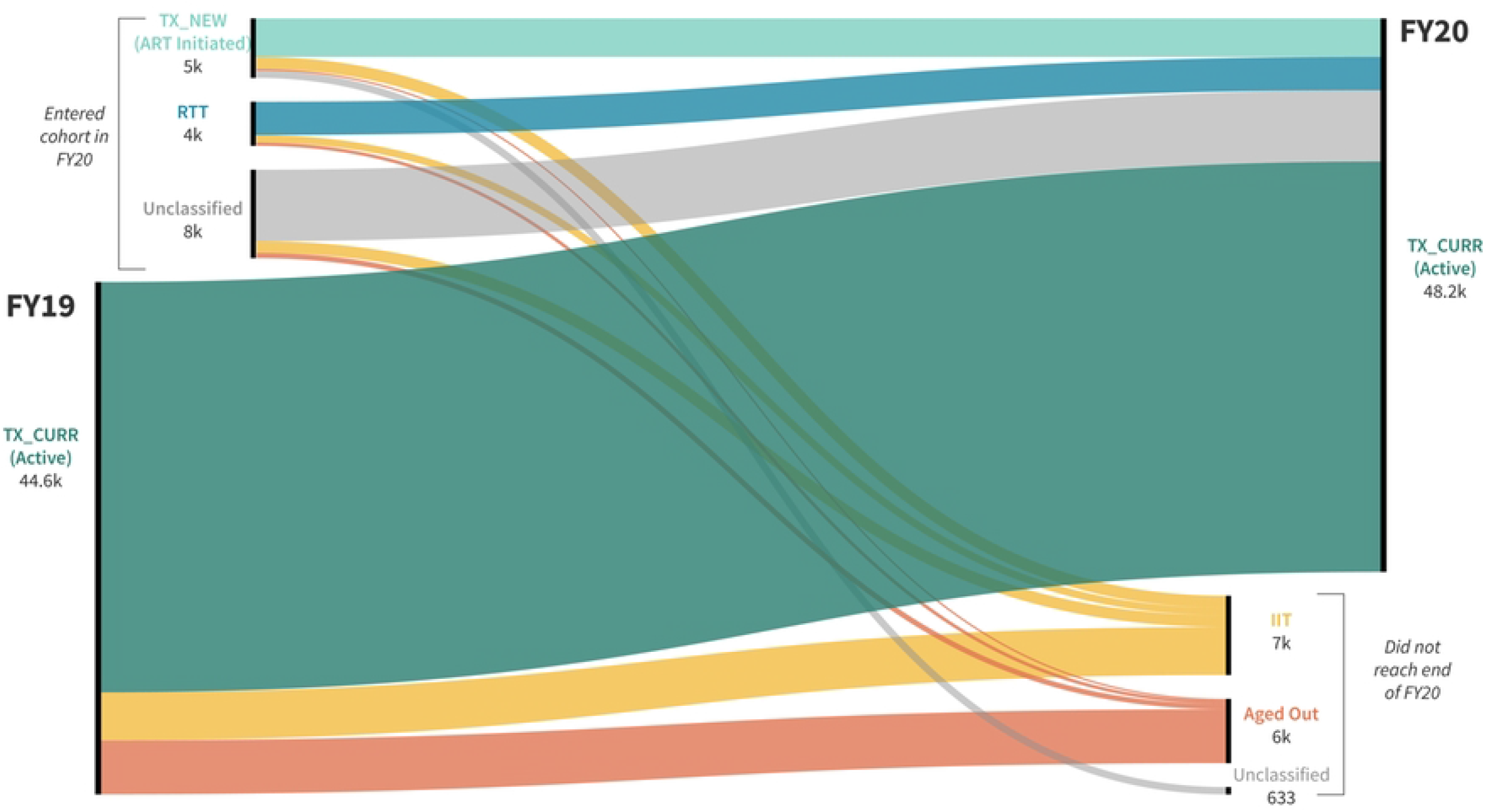
Tracking treatment outcomes among C/ALHIV in Kenya – FY2019 - FY2020.

**Fig 2.**
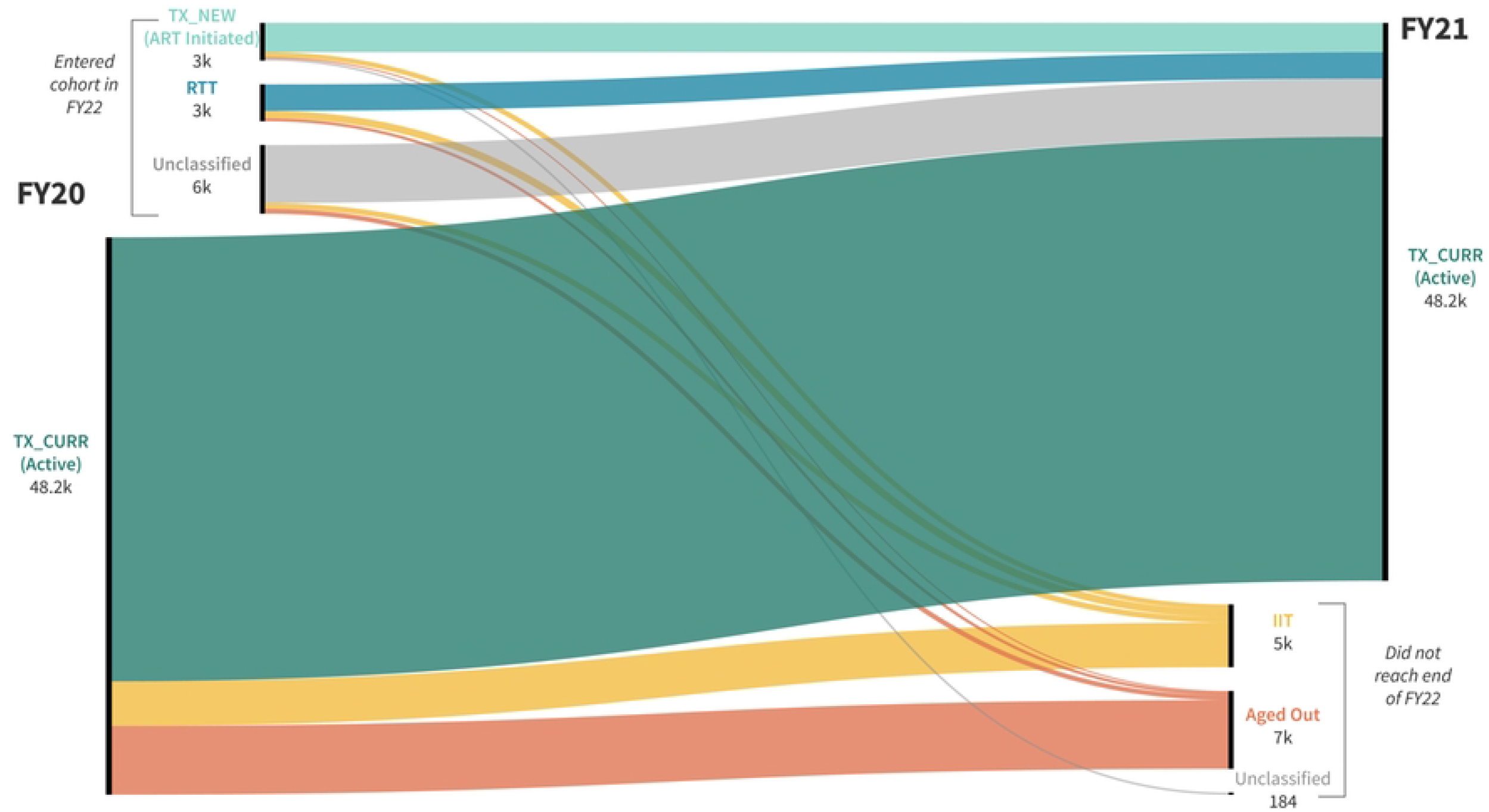
Tracking treatment outcomes among C/ALHIV in Kenya – FY2020 - FY2021.

**Fig 3.**
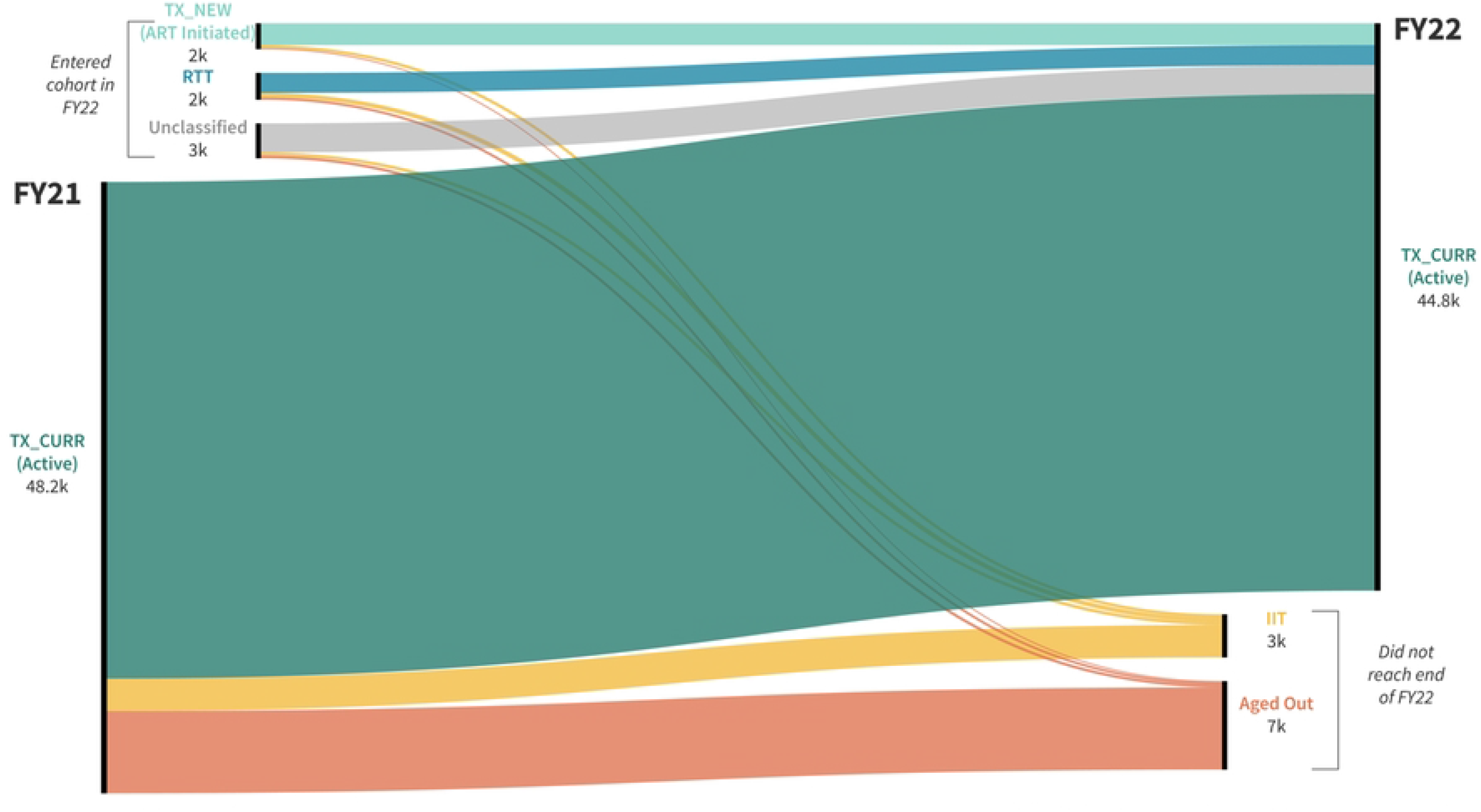
Tracking treatment outcomes among C/ALHIV in Kenya – FY2021 - FY2022.

## Discussion

Normal aging-out of the <15-year-old cohort comprises a significant source of cohort “loss” among C/ALHIV continuing ART for each FY from 2020-2022 and is greater than IIT in both FY2021 and FY2022. Impact of aging-out is not routinely captured in programmatic PEPFAR data and requires individual-level data analysis for identification. In this analysis, impact of aging-out results in CoT estimates that underestimate actual CoT for C/ALHIV. Additionally, C/ALHIV who experienced IIT decreased each FY under study, perhaps due to successful programmatic interventions such as Operation Triple Zero - a customized adolescent-focused service delivery model for “Zero viral load, Zero missed appointments, and Zero missed drugs”, implemented around the same timeframe [17].

When a C/ALHIV is not retained on ART, understanding IIT root causes allows for tailored CoT interventions that improve clinical outcomes. Programs can then provide targeted support to C/ALHIV on ART who have aged out of the cohort, but remain on treatment, to ensure successful transition to adulthood. Focused, person-centered support is particularly important as demonstrated through a recent systematic review that indicated wide variation (37 to 97.4%) of CoT rates among ALHIV following transition to adult services [18].

Based on our analyses that allowed for accurate estimation of CoT, future directions would include analyzing individual-level clinical outcomes among subsets of C/ALHIV who experienced IIT. Analysis of IIT root causes among C/ALHIV at clinical sites will shed light on specific reasons for ART interruption and provide actionable data to optimize program implementation. Additionally, individual-level analyses are needed to verify if adolescents who aged out of pediatric cohorts (0-14 years) have successfully transitioned to adult cohorts (15+ years). Kenya’s National Unique Patient Identifier program utilizes birth certificate registration numbers to provide unique identifier numbers to C/ALHIV. Future Kenyan analyses can utilize these identifiers to deduplicate and track outcomes of C/ALHIV who experience IIT.

This analysis has some limitations. Firstly, it excludes Kenyan treatment sites that utilize paper records and sites in seven low-HIV burden counties potentially affecting generalizability of findings. As of Jan 2022, 85% (1,046,761) of all clients on ART were seen at an EMR-site. Secondly, annual analysis may mask short-term trends. And lastly, the study population focuses on those with documented interruptions, omitting transfers and deaths.

“Aging Out” analyses are now easier to perform - EMRs are more prevalent; access to patient-level data has increased; standardized protocols for loss analysis are refined; and PEPFAR guidance is now explicit in calling for these analyses. However, performing these analyses separately for each age-band and implementing partner remains burdensome. Ideally these analyses would be performed through unified host country data systems that house patient-level data reported directly to Ministries of Health (MoH), and MoH partners. This would permit speedy identification of gaps in CoT, adjusting for losses and gains due to normal aging-out. Ultimately, by improving individual-level clinical outcomes for C/ALHIV, these analyses bolster long-term sustainability of host country HIV treatment programs.

## Conclusions

This analysis suggests that normal aging-out results in underestimation of HIV treatment continuity for C/ALHIV. Routine aging-out analyses can inform programs on their true rates of IIT, as they work to achieve epidemic control among C/ALHIV.

## Data Availability

The data that support the findings of this study are openly available in the Kenya National Data Warehouse at https://dwh.nascop.org/#/

## Acknowledgements

We thank Nassir Kashmiri, formerly with Management and Development for Health (MDH), Tanzania, and Roland Van de Ven, USAID Afya Yangu Northern, Tanzania, for sharing unpublished data with the study authors, and Ohvia Muraleetharan who worked on the initial abstract for this paper.

